# Deploying Local Large Language Models for Automated Article Screening in Scientific Literature Reviews

**DOI:** 10.64898/2026.10.05.26364746

**Authors:** Ling-Hsuan Lao, Hongruyu Chen, William Sutcliffe, Henock G. Yebyo, Nicola Serra, Milo Alan Puhan

## Abstract

**Background:** Large language models (LLMs) show promise in scientific literature reviews, but cloud-based proprietary models like GPT-4 raise concerns around privacy, copyright, and licensed article access. Using a scoping review on AI applications for treatment effect estimation as a use case, this study evaluated local open-source LLMs for automated article screening in scientific literature reviews and developed a practical workflow for their implementation.

**Methods:** We extracted a sample dataset of 300 records from an ongoing scoping review on machine learning methods for treatment effect estimation using real-world medical data. We divided the dataset into training (n = 200) and testing (n = 100) datasets. We evaluated 11 locally deployed open-source LLMs from five model families (GPT-OSS, Gemma, Qwen, Mistral, Llama) for both abstract screening and full-text screening. Full-text screening was done using text converted by two PDF-to-text conversion tools: Docling and PyMuPDF4LLM. Performance was assessed using raw accuracy with 95% Wilson’s confidence interval, relaxed accuracy, Cohen’s kappa, and runtime. During workflow development, prompt engineering, output extraction strategies, and generation settings were iteratively optimized using the training dataset.

**Results:** GPT-OSS-20B achieved the highest raw accuracy for title and abstract screening (0.890 on the training set and 0.850 on the test set), followed closely by Gemma-4-31B-it (0.875 on the training set and 0.830 on the test set) and Gemma-4-26B-A4B-it (0.810 on the training set and 0.790 on the test set). In full-text screening, slightly higher raw accuracy and reduced false-positive rates were observed, with GPT-OSS-20B and Gemma-4 variants consistently demonstrating strong performance. Only minor variations were found between the two PDF-to-text conversion methods evaluated, though runtime varied substantially across models. Based on these findings, we compiled a set of practical recommendations for local LLM-assisted article screening.

**Conclusions:** Local LLMs can effectively support literature screening when integrated into a carefully designed workflow. Recent generations of open-source local models achieved performance comparable to human reviewers and proprietary LLMs. Local deployment also provides advantages in data privacy, copyright compliance, customization, and cost-efficiency. While human oversight remains essential, the proposed workflow demonstrated that local LLMs represent a practical and reproducible approach for accelerating evidence synthesis.

## Introduction

Scientific literature reviews, including systematic reviews and scoping reviews, play an important role in synthesizing evidence across studies, identifying research gaps, and characterizing a given topic [1–3]. However, such reviews involve multiple predominantly manual steps, making the process labor-intensive and time-consuming [4–6], especially as the volume of literature continues to grow [7]. Borah et al. [8] estimated that completing a review takes an average of 67 weeks, and involves a mean of five authors. Among the various review stages, article screening is particularly important because review quality of reviews largely relies on comprehensive database searches and rigorous selection of eligible studies [9]. Article screening is also one of the most time-intensive steps in the systematic review process [10]. For example, in a study from Polanin et al. [11], research staff spent 189 days screening 29,846 abstracts independently. In addition, large screening workloads can increase the risk of human error and reviewer fatigue, potentially affecting screening decisions [12].

Given these challenges, there has been growing interest in using natural language processing (NLP) and machine learning methods to support article screening in systematic reviews [13, 14]. Widely used tools include Rayyan and ASReview. However, these tools remain largely reviewer-in-the-loop systems focused on title and abstract prioritization, with limited capacity for flexible, criterion-based full-text assessment. They primarily function as decision-support tools rather than independently making screening decisions [15, 16]. More recently, the emergence of Large Language Models (LLMs), have generated growing interest in the potential for more autonomous literature screening tasks [4].

LLMs have shown strong capabilities across a range of natural language processing tasks and are increasingly being applied in academic research [4]. Trained on massive text datasets, LLMs are particularly well-suited to intensive text-based tasks such as summarization, information extraction, translation, and question answering. And they can often perform effectively without extensive domain-specific training. These characteristics highlight their potential to streamline the systematic review process [17, 18]. Recent LLMs such as GPT, Gemini and Claude have attracted growing interest in evidence synthesis tasks, especially for citation screening in systematic reviews [19, 20]. Although current systems are not yet fully autonomous and show variable performance, studies have demonstrated that well-developed LLM-based workflows can achieve performance comparable to human reviewers while substantially improving screening efficiency [5].

To date, most studies evaluating LLMs in article screening for scientific literature reviews have focused on proprietary models, particularly GPT-3.5 and GPT-4 [21]. Their widespread adoption is likely driven by their strong performance in language understanding and reasoning tasks [22, 23]. Despite these advantages, proprietary LLMs have important limitations. Most require users to transmit data through external APIs, raising concerns about data privacy, intellectual property [24], and compliance with licensing restrictions when copyrighted scholarly content is processed by third-party services [25]. Limited transparency and frequent model updates may reduce reproducibility and make model behavior difficult to understand or validate [22, 26]. Consequently, there has been growing interest in open-source and locally deployable LLMs. Local LLMs can be deployed within institutional infrastructure, offering lower operating costs, greater customizability, improved transparency, and greater control over data processing and security. These features may be particularly valuable for article screening workflows involving copyrighted or sensitive research materials [23, 27, 28]. However, the performance of open-source LLMs in article screening remains less well established than that of proprietary models.

To address this gap, this study evaluated the performance of 11 open-source Local LLMs on automated abstract screening and full-text screening for scientific literature reviews. We assessed their utility using a scoping review on AI methods for treatment effect estimation in medical research. Based on the findings, we provide practical recommendations for researchers seeking cost-effective, secured, and locally controlled LLM-assisted screening workflows for scientific literature reviews.

## Methods

### Study Design

We evaluated the performance of 11 locally deployed, open-source LLMs for automated article screening in scientific literature reviews using a sample of 300 articles derived from a large-scale database search. The dataset was divided into a training set (*n* = 200) and a held-out test set (*n* = 100). The training set was used for prompt engineering, output extraction and parameter tuning, while the test set was used to independently assess model performance using several quantitative metrics. The screening workflow consisted of two stages: title and abstract screening followed by full-text screening. During title and abstract screening, studies were classified into three categories: “Relevant”, “Not Relevant” or “Uncertain”. Hereafter, we use abstract screening to indicate title and abstract screening for simplicity. Full-text screening used binary classification (Relevant or Not Relevant), assessing the models’ ability in evaluating complete articles against eligibility criteria. A schematic overview of the workflow is presented in Figure 1.

**Figure 1.**
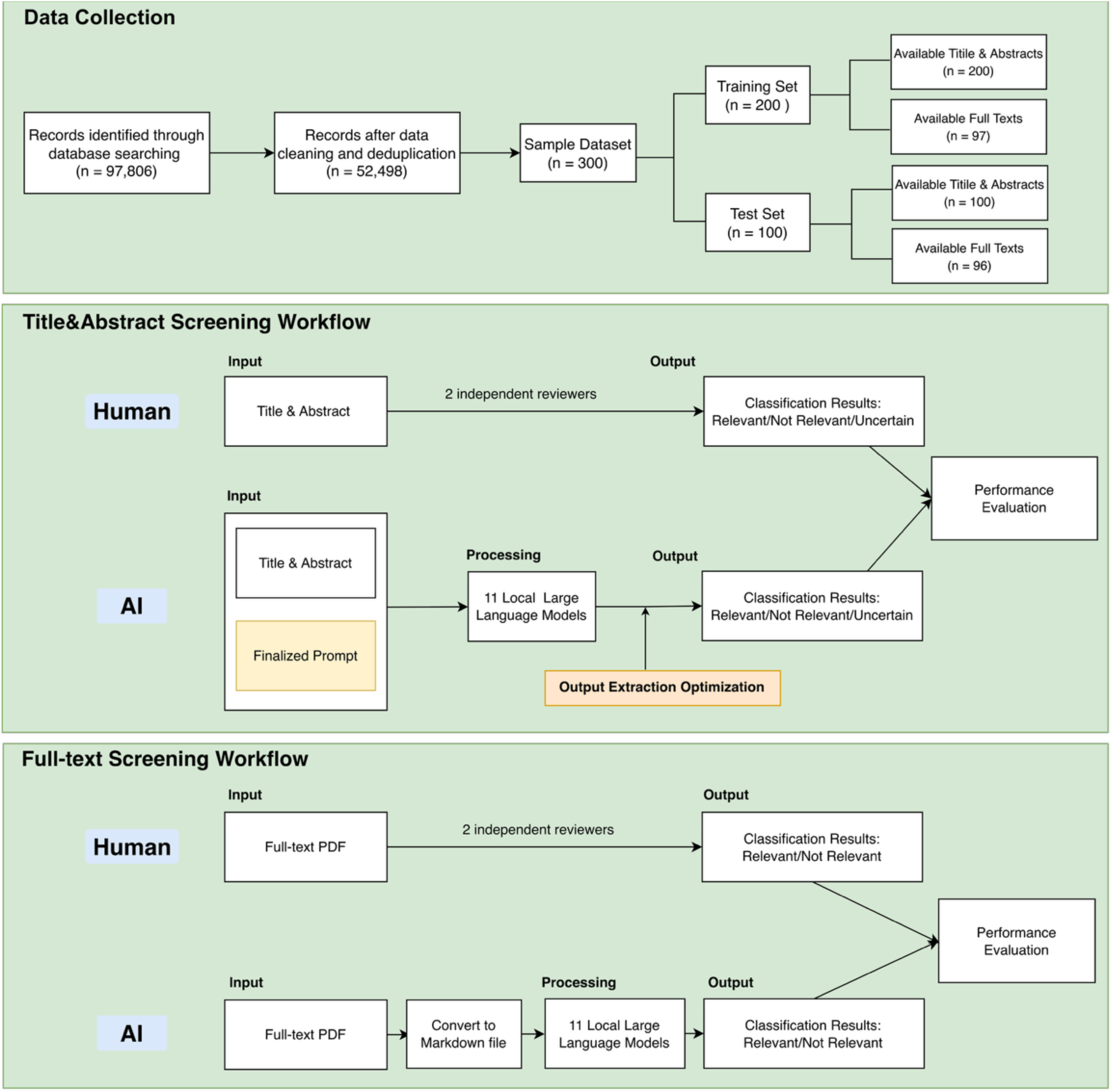
Workflow of scientific literature screening. Following database searching, data cleaning, and deduplication, a sample dataset of 300 records was constructed and randomly divided into a training set (n = 200) and a testing set (n = 100). The training set was used to optimize prompt design and model-specific output extraction strategies (see Figure 2), whereas the optimized workflow was subsequently applied to the independent testing set for performance evaluation. Two human reviewers performed title and abstract screening as well was full-text screening independently, and model classifications were compared with human classifications.

### Literature Search and Sample Dataset

This study was conducted as part of our ongoing broader scoping review aimed at identifying research using artificial intelligence (AI) methods to estimate treatment effects in real-world medical applications. With the assistance of an experienced medical librarian, we performed a systematic search across six bibliographic databases: Embase, Medline, Cochrane, Web of Science, Scopus, and the ACM Digital Library. Detailed search strings are provided in Supplementary Materials S1. The search was completed on April 30, 2025.

Following deduplication and data cleaning of the search results, we selected a random sample of 300 articles. Inclusion criteria were articles published in the English language that applied AI methods to treatment effect estimation using real-world medical data. We excluded studies that met any of the following criteria: (1) non-causal objectives (e.g., risk factor identification or outcome prediction); (2) relied on traditional statistical methods rather than AI; (3) lacked treatment effect assessment; or (4) used only simulated or synthetic datasets.

Each entry in the dataset included the paper’s metadata, a human classification (Relevant, Not Relevant, or Uncertain), and the corresponding reasoning explaining the classification. To ensure a robust evaluation, we purposively sampled to maintain a balanced distribution across all screening classifications. To establish a reference standard, two reviewers independently screened all 300 articles manually. Any discrepancies were resolved through consensus discussion. The resulting consensus classifications served as the ground truth for model evaluation.

### LLM-based Classification Framework

We evaluated 11 open-source LLMs that are publicly available on Hugging Face, including GPT-OSS-20B, Gemma-3-12b-it, Gemma-3-27b-it, Gemma-4-26B-A4B-it, Gemma-4-31B-it, Qwen3-14B, Qwen3-4B-Instruct-2507, Ministral-3-14B-Instruct-2512, Ministral-3-3B-Instruct-2512, Ministral-3-14B-Reasoning-2512, and Llama-3.1-8B-Instruct. Experiments were conducted on a server equipped with four NVIDIA L40S GPUs (48 GB VRAM each) CUDA 12.8, and two AMD EPYC 9654 CPUs (96 cores per socket, 192 cores total, 384 threads). Models were deployed using automatic device mapping to enable multi-GPU inference. For each model, we recorded the commit hash to ensure reproducibility [29].

We developed an automated classification framework in which LLM inputs consisted of structured metadata (e.g., title, authors, and abstract) and fine-tuned prompts. Model outputs were parsed into predefined classification categories: ternary classification (Relevant, Not Relevant, Uncertain) for abstract screening, and binary classification (Relevant, Not Relevant) for full-text screening, aligning with the human screening process. The LLM inference was performed independently of the human classifications to minimize potential bias. Local deployment of these models required sufficient VRAM capacity. Selection of the models considered both hardware constraints and model diversity, encompassing both reasoning-optimized models and general-purpose instruction models. The selection also reflected state of art performance within the 4-32B parameter range given the GPU VRAM constraints. This variety allowed us to assess how different model architectures handle scientific literature screening. A summary of model characteristics is provided in Supplementary Materials S2.

### Prompt Engineering and Output Extraction

A prompt, an instruction that guides LLMs to generate task-specific outputs, was developed using the training dataset through iterative human refinement based on observed model misclassifications. The final prompt instructed the LLMs to classify studies according to their relevance to AI methods for treatment effect estimation using real-world medical data. The prompt consisted of three components:

1. Clear task definition: The prompt explicitly instructed the LLM to perform screening solely based on the provided metadata.
2. Criteria and classification rules: The prompt defined three criteria for classification: use of AI methods, estimation of treatment effects, and use of real-world medical data. Classification was defined using logical rules: studies were labelled as “Relevant” if all three criteria were clearly satisfied, “Not Relevant” if any one of the three criteria was clearly not satisfied, and “Uncertain” if at least one criterion was unclear while the remaining criteria were satisfied.
3. Output specification: LLMs were required to return the classification for each metadata entry restricted to “Relevant,” “Not Relevant,” or “Uncertain”.

We adopted a zero-shot learning approach [30], in which no example studies were included in the prompt. In addition, output extraction strategies were tailored to each LLM to accommodate differences in response formats across models. We also tuned the output token limit to balance computational efficiency and classification accuracy (See Figure 2). Subsequently, we applied the optimized prompt to the test set to evaluate model performance. The prompt used in this study is provided in Supplementary Material S3. The prompt for the full-text screening phase was identical to the abstract screening prompt, with the exception that the “Uncertain” classification label was removed.

**Figure 2.**
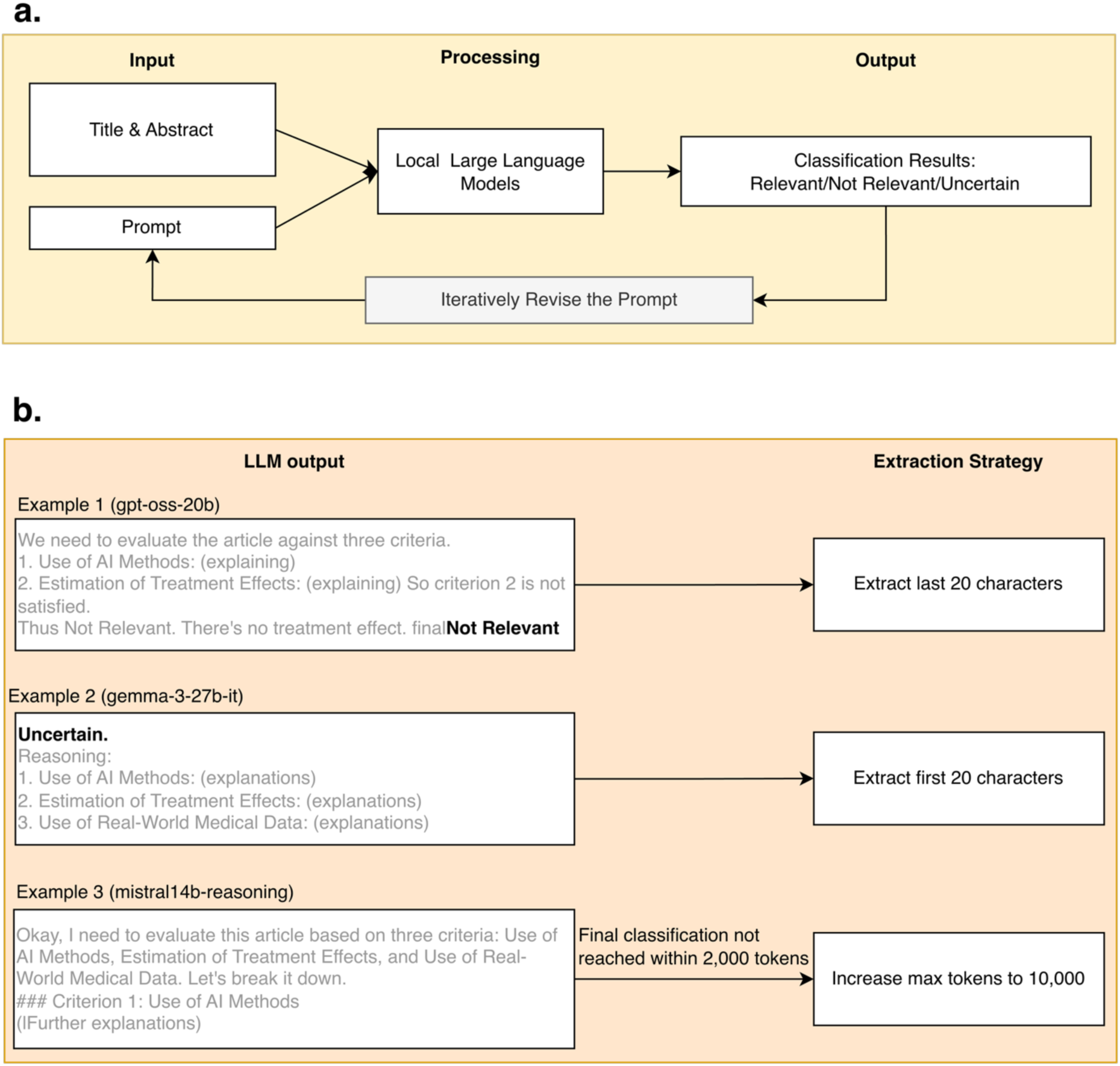
Optimization procedures performed on the training set. **a**. Prompt optimization. The training set was used to iteratively refine prompts based on model outputs and observed classification errors. b. Output extraction optimization. The training set was also used to develop output extraction strategies for each model according to each model’s response format, including character-based extraction and adjustment of the maximum token limit.

### Abstract Screening

In the first stage, we conducted LLM screening using only the extracted titles and abstracts. These data were organized in a structured tabular format. For each iteration, the title and abstract of one record were provided as input to the locally deployed LLMs, together with the optimized prompt developed. Based on this information, each LLM independently assessed the relevance of each record without access to the full texts. The models were instructed to assign one of three labels: “Relevant,” “Not Relevant,” or “Uncertain”. The inference process was conducted in a fully automated and blinded manner. The resulting classifications were stored in a structured format for downstream quantitative evaluation. During inference, to reduce output variability and improve reproducibility, greedy decoding `(do_sample = FALSE`) was used, thereby the highest-probability token was selected at each decoding step, yielding deterministic outputs. This stage reflects the initial screening step commonly used in systematic reviews, where decisions are made based on limited information.

### Full-text Screening

In the second stage, we evaluated LLM performance for full-text screening. Because key eligibility information is often absent or ambiguous in abstracts, we converted each paper’s full-text PDF to machine-readable Markdown so that the models could classify each study using its complete content. We compared two open-source PDF-to-Markdown converters, Docling [31] and PyMuPDF4LLM [32]. Both preserved the textual content and reading order well across heterogeneous journal layouts, but they handled structural elements differently. PyMuPDF4LLM is a rule-based extractor that preserves in-text hyperlinks but reproduces the PDF’s physical layout: body text is hard-wrapped at the original line breaks, running headers and page footers (e.g. journal citation lines and “Page N of M”) are retained within the article body, figures are dropped without markers, and tabular content is emitted as space-separated text that is rarely recovered as a structured table. Docling instead applies document-layout and table-structure models, reflowing paragraphs into continuous text, removing repeated page structure, inserting explicit figure placeholders, and reconstructing tables as valid Markdown pipe tables in most documents. To measure the downstream effect of these differences, we converted all sample papers with both tools and ran the 11 local LLMs on each version.

Human reviewers re-assessed each paper against the full text and labelled it “Relevant” or “Not Relevant”; these labels served as ground truth. To accommodate hardware limitations, each paper’s text was truncated to a per-model character budget (20,000–40,000 characters). While this truncation may have excluded relevant content from longer manuscripts, it also served the practical purpose of removing unnecessary text to accelerate screening times.

### Evaluation Metrics

We assessed two accuracy metrics. Raw accuracy was defined as the proportion of exact matches between the model and human labels. Relaxed accuracy counted every prediction as correct except false negatives—cases where the model assigns “Not Relevant” to a paper the human reviewers judged eligible (i.e., Relevant or Uncertain). This reflects the priority in systematic-review screening to maximize recall and avoid discarding potentially eligible studies. For raw accuracy, we report 95% Wilson score confidence intervals. Chance-corrected agreement between each model and the human labels was quantified with Cohen’s kappa (κ). Finally, we recorded the runtime of each screening run to compare computational cost across models (and, at the full-text stage, between the two conversion tools). The same set of metrics was applied to both the abstract and full-text screening stages, comparing each model’s classifications against the human ground truth. All metrics were computed separately for the training and test sets, and—at the full-text stage—separately for the Docling and PyMuPDF4LLM conversions.

## Results

### Sample Dataset

As part of the scoping review of AI use for treatment effect estimation, we identified 97,806 records across six databases (Figure 1). Following data cleaning and deduplication, 52,498 unique records remained. From these records, a sample of 300 articles was selected for LLM screening. Based on title and abstract, human reviewers labelled 122 articles (40.7%) Relevant, 34 (11.3%) Uncertain, and 144 (48.0%) Not Relevant. The distribution was similar across the two sets: in the training set (*n* = 200), 76 Relevant, 26 Uncertain, and 98 Not Relevant; in the test set (*n* = 100), 46 Relevant, 8 Uncertain, and 46 Not Relevant. At the full-text stage, the full text could be retrieved and converted for 293 of the 300 articles (the remaining 7 were unavailable and excluded from this stage). Overall, 136 articles (46.4%) were classified Relevant and 157 (53.6%) Not Relevant. By split, the training set (*n* = 197) contained 92 Relevant and 105 Not Relevant, and similarly the test set (*n* = 96) contained 44 Relevant and 52 Not Relevant.

### Abstract Screening

Considerable variation in performance was observed across models (Table 1). On the training set, raw accuracy ranged from 0.505 to 0.890. GPT-OSS-20B achieved the highest raw accuracy (0.890), followed by Gemma-4-31B-it (0.875), Gemma-4-26B-A4B-it (0.810), and Qwen3-14B (0.760). Smaller models generally achieved lower raw accuracy, including Gemma-3-12B-it (0.505) and Qwen3-4B-Instruct-2507 (0.565). Similar trends were observed on the test set, where GPT-OSS-20B again achieved the highest raw accuracy (0.850), followed by Gemma-4-31B-it (0.830), Gemma-4-26B-A4B-it (0.790), Ministral-3-14B-Reasoning-2512 (0.760) and Qwen3-14B (0.740). Across all 11 models, training and test accuracy differed by less than 0.10. Relaxed accuracy was substantially higher than raw accuracy across all evaluated models. On the training set, relaxed accuracy ranged from 0.885 to 0.995, while on the test set, it ranged from 0.900 to 1.000. This reflects that most disagreements were over-inclusions rather than missed relevant articles.

**Table 1.**
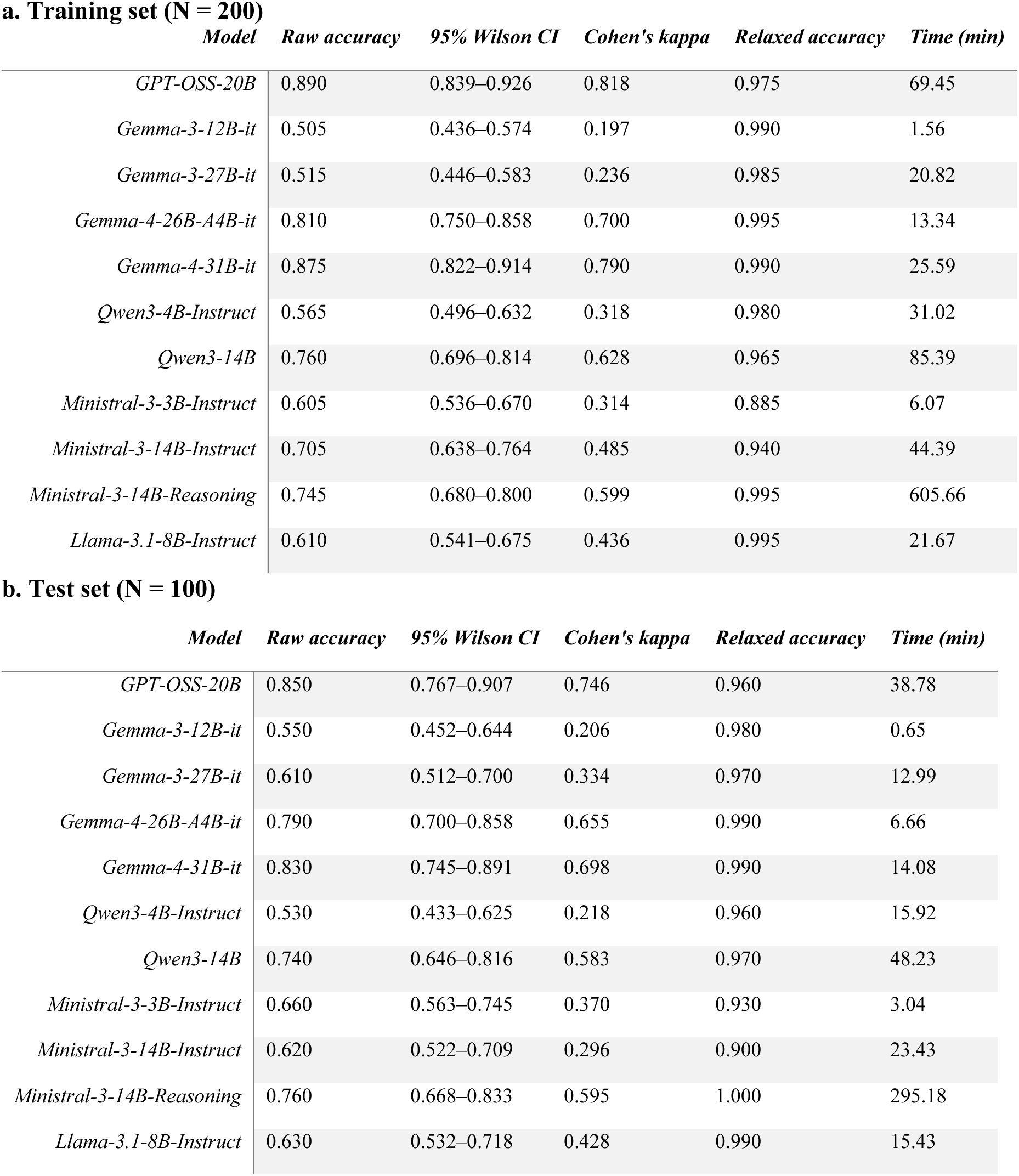
Title and abstract screening performance on training and test dataset across 11 local Large Language Models. **a**. Results on training set. **b**. Results on test set.

**a. Training set (N = 200)**
| *Model* | *Raw accuracy* | *95% Wilson CI* | *Cohen's kappa* | *Relaxed accuracy* | *Time (min)* |
| --- | --- | --- | --- | --- | --- |
| *GPT-OSS-20B* | 0.890 | 0.839–0.926 | 0.818 | 0.975 | 69.45 |
| *Gemma-3-12B-it* | 0.505 | 0.436–0.574 | 0.197 | 0.990 | 1.56 |
| *Gemma-3-27B-it* | 0.515 | 0.446–0.583 | 0.236 | 0.985 | 20.82 |
| *Gemma-4-26B-A4B-it* | 0.810 | 0.750–0.858 | 0.700 | 0.995 | 13.34 |
| *Gemma-4-31B-it* | 0.875 | 0.822–0.914 | 0.790 | 0.990 | 25.59 |
| *Qwen3-4B-Instruct* | 0.565 | 0.496–0.632 | 0.318 | 0.980 | 31.02 |
| *Qwen3-14B* | 0.760 | 0.696–0.814 | 0.628 | 0.965 | 85.39 |
| *Ministral-3-3B-Instruct* | 0.605 | 0.536–0.670 | 0.314 | 0.885 | 6.07 |
| *Ministral-3-14B-Instruct* | 0.705 | 0.638–0.764 | 0.485 | 0.940 | 44.39 |
| *Ministral-3-14B-Reasoning* | 0.745 | 0.680–0.800 | 0.599 | 0.995 | 605.66 |
| *Llama-3.1-8B-Instruct* | 0.610 | 0.541–0.675 | 0.436 | 0.995 | 21.67 |

**b. Test set (N = 100)**
| *Model* | *Raw accuracy* | *95% Wilson CI* | *Cohen's kappa* | *Relaxed accuracy* | *Time (min)* |
| --- | --- | --- | --- | --- | --- |
| *GPT-OSS-20B* | 0.850 | 0.767–0.907 | 0.746 | 0.960 | 38.78 |
| *Gemma-3-12B-it* | 0.550 | 0.452–0.644 | 0.206 | 0.980 | 0.65 |
| *Gemma-3-27B-it* | 0.610 | 0.512–0.700 | 0.334 | 0.970 | 12.99 |
| *Gemma-4-26B-A4B-it* | 0.790 | 0.700–0.858 | 0.655 | 0.990 | 6.66 |
| *Gemma-4-31B-it* | 0.830 | 0.745–0.891 | 0.698 | 0.990 | 14.08 |
| *Qwen3-4B-Instruct* | 0.530 | 0.433–0.625 | 0.218 | 0.960 | 15.92 |
| *Qwen3-14B* | 0.740 | 0.646–0.816 | 0.583 | 0.970 | 48.23 |
| *Ministral-3-3B-Instruct* | 0.660 | 0.563–0.745 | 0.370 | 0.930 | 3.04 |
| *Ministral-3-14B-Instruct* | 0.620 | 0.522–0.709 | 0.296 | 0.900 | 23.43 |
| *Ministral-3-14B-Reasoning* | 0.760 | 0.668–0.833 | 0.595 | 1.000 | 295.18 |
| *Llama-3.1-8B-Instruct* | 0.630 | 0.532–0.718 | 0.428 | 0.990 | 15.43 |

Within model families, the newer Gemma-4 models substantially outperformed the earlier Gemma-3 models. On the test set, raw accuracy rose from 0.55–0.61 for Gemma 3 models to 0.79–0.83 for Gemma 4 models. Within the Qwen family, Qwen3-14B outperformed Qwen3-4B-Instruct-2507 on both the training set (0.760 vs. 0.565) and the test set (0.740 vs. 0.530). Within the Mistral family, the reasoning variant achieved the highest raw accuracy on both the training set (0.745) and test set (0.760), though this came at substantially longer inference times than the other Mistral models.

The 95% Wilson score confidence intervals spanned approximately ±0.04–0.07 on the training set and ±0.07–0.10 on the test set. These intervals distinguished the strongest models from the weakest but not the top performers from one another; on the training set, GPT-OSS-20B (0.890, 95% CI 0.839–0.926) and Gemma-4-31B-it (0.875, 95% CI 0.822–0.914) had heavily overlapping intervals, whereas both clearly exceeded Qwen3-14B (0.760, 95% CI 0.696–0.814) and the Gemma-3 models (≈0.51, upper bounds < 0.59). Chance-corrected agreement with the human labels (κ) tracked the accuracy ranking, suggesting that the differences were not simply an artifact of class imbalance: on the training set κ ranged from 0.20 (Gemma-3-12B-it) to 0.82 (GPT-OSS-20B), with GPT-OSS-20B and the Gemma-4 models reaching substantial-to-almost-perfect agreement (κ ≈ 0.70–0.82), the mid-tier models moderate-to-substantial agreement (Qwen3-14B κ = 0.63; Ministral-3-14B-Reasoning κ = 0.60), and the Gemma-3 and Qwen3-4B models only slight-to-fair agreement (κ ≤ 0.32). The same pattern was observed on the test set (κ from 0.21 to 0.75).

Inference time varied by more than two orders of magnitude. On the training set, it ranged from 1.56 minutes (Gemma-3-12B-it) to 605.66 minutes (10.09 hours) for Ministral-3-14B-Reasoning-2512. On the test set, the runtime ranged from 0.65 minutes (Gemma-3-12B-it) to 295.18 minutes (4.92 hours) for Ministral-3-14B-Reasoning-2512. Among the higher-performing models, GPT-OSS-20B required 69.45 and 38.78 minutes on the training and test sets, whereas Gemma-4-31B-it only 25.59 and 14.08 minutes, respectively.

Disabling sampling (`do_sample=False`) resulted in more deterministic model outputs across runs. In terms of performance, both raw accuracy and relaxed accuracy remained largely unchanged. An exception was observed for the Ministral-3-14B-Reasoning-2512 model, where disabling sampling was associated with unstable outputs. Consequently, this parameter setting was not applied to the model and majority voting across three runs was used instead.

### Full-text Screening

Performance trends were broadly consistent across the two converters (Table 2). On the training set, GPT-OSS-20B achieved the highest raw accuracy under both Docling (0.899) and PyMuPDF4LLM (0.883). Gemma-4-31B-it and Gemma-4-26B-A4B-it followed closely (0.888 and 0.873 with Docling, 0.883 and 0.878 with PyMuPDF4LLM, respectively). Qwen3-14B also demonstrated strong performance (0.863 with Docling and 0.853 with PyMuPDF4LLM). As in the abstract stage, the Gemma-3 models performed substantially worse, with raw accuracy ranging from 0.553 to 0.594.

**Table 2.** Full-text screening performance on training and test dataset across 11 local Large Language Models. **a**. Results on training set based on Docling. **b**. Results on training set based on PyMuPDF4LLM. **c.** Results on test set based on Docling. **d**. Results on test set based on PyMuPDF4LLM.

**a. Training set — Docling (N = 197)**
| *Model* | *Raw accuracy* | *95% Wilson CI* | *Cohen's kappa* | *Relaxed accuracy* |
| --- | --- | --- | --- | --- |
| *GPT-OSS-20B* | 0.898 | 0.848–0.933 | 0.795 | 0.939 |
| *Gemma-3-12B-it* | 0.553 | 0.483–0.621 | 0.090 | 0.726 |
| *Gemma-3-27B-it* | 0.569 | 0.499–0.636 | 0.085 | 0.584 |
| *Gemma-4-26B-A4B-it* | 0.873 | 0.819–0.912 | 0.748 | 0.980 |
| *Gemma-4-31B-it* | 0.888 | 0.837–0.925 | 0.777 | 0.965 |
| *Qwen3-4B-Instruct* | 0.665 | 0.597–0.727 | 0.345 | 0.939 |
| *Qwen3-14B* | 0.863 | 0.808–0.904 | 0.726 | 0.949 |
| *Ministral-3-3B-Instruct* | 0.706 | 0.638–0.765 | 0.408 | 0.848 |
| *Ministral-3-14B-Instruct* | 0.807 | 0.746–0.856 | 0.616 | 0.939 |
| *Ministral-3-14B-Reasoning* | 0.756 | 0.692–0.811 | 0.527 | 0.975 |
| *Llama-3.1-8B-Instruct* | 0.634 | 0.565–0.699 | 0.293 | 0.965 |

**b. Training set — PyMuPDF4LLM (N = 197)**
| *Model* | *Raw accuracy* | *95% Wilson CI* | *Cohen's kappa* | *Relaxed accuracy* |
| --- | --- | --- | --- | --- |
| *GPT-OSS-20B* | 0.883 | 0.831–0.921 | 0.764 | 0.924 |
| *Gemma-3-12B-it* | 0.594 | 0.524–0.660 | 0.181 | 0.782 |
| *Gemma-3-27B-it* | 0.584 | 0.514–0.650 | 0.119 | 0.599 |
| *Gemma-4-26B-A4B-it* | 0.878 | 0.825–0.917 | 0.758 | 0.975 |
| *Gemma-4-31B-it* | 0.883 | 0.831–0.921 | 0.766 | 0.954 |
| *Qwen3-4B-Instruct* | 0.685 | 0.617–0.746 | 0.387 | 0.959 |
| *Qwen3-14B* | 0.853 | 0.797–0.895 | 0.704 | 0.924 |
| *Ministral-3-3B-Instruct* | 0.726 | 0.660–0.783 | 0.459 | 0.914 |
| *Ministral-3-14B-Instruct* | 0.833 | 0.774–0.878 | 0.665 | 0.939 |
| *Ministral-3-14B-Reasoning* | 0.756 | 0.692–0.811 | 0.537 | 0.985 |
| *Llama-3.1-8B-Instruct* | 0.624 | 0.555–0.689 | 0.278 | 0.980 |

**c. Test set — Docling (N = 96)**
| *Model* | *Raw accuracy* | *95% Wilson CI* | *Cohen's kappa* | *Relaxed accuracy* |
| --- | --- | --- | --- | --- |
| *GPT-OSS-20B* | 0.833 | 0.746–0.895 | 0.667 | 0.938 |
| *Gemma-3-12B-it* | 0.625 | 0.525–0.715 | 0.229 | 0.750 |
| *Gemma-3-27B-it* | 0.573 | 0.473–0.667 | 0.073 | 0.573 |
| *Gemma-4-26B-A4B-it* | 0.792 | 0.700–0.861 | 0.592 | 0.979 |
| *Gemma-4-31B-it* | 0.812 | 0.723–0.878 | 0.629 | 0.958 |
| *Qwen3-4B-Instruct* | 0.646 | 0.546–0.734 | 0.315 | 0.948 |
| *Qwen3-14B* | 0.792 | 0.700–0.861 | 0.589 | 0.958 |
| *Ministral-3-3B-Instruct* | 0.729 | 0.633–0.808 | 0.455 | 0.865 |
| *Ministral-3-14B-Instruct* | 0.708 | 0.611–0.790 | 0.419 | 0.885 |
| *Ministral-3-14B-Reasoning* | 0.708 | 0.611–0.790 | 0.448 | 0.990 |
| *Llama-3.1-8B-Instruct* | 0.594 | 0.494–0.686 | 0.219 | 0.938 |

**d. Test set — PyMuPDF4LLM (N = 96)**
| *Model* | *Raw accuracy* | *95% Wilson CI* | *Cohen's kappa* | *Relaxed accuracy* |
| --- | --- | --- | --- | --- |
| *GPT-OSS-20B* | 0.885 | 0.806–0.935 | 0.771 | 0.969 |
| *Gemma-3-12B-it* | 0.562 | 0.463–0.657 | 0.100 | 0.719 |
| *Gemma-3-27B-it* | 0.615 | 0.515–0.706 | 0.179 | 0.646 |
| *Gemma-4-26B-A4B-it* | 0.823 | 0.735–0.886 | 0.651 | 0.979 |
| *Gemma-4-31B-it* | 0.812 | 0.723–0.878 | 0.629 | 0.958 |
| *Qwen3-4B-Instruct* | 0.667 | 0.568–0.753 | 0.349 | 0.927 |
| *Qwen3-14B* | 0.792 | 0.700–0.861 | 0.585 | 0.927 |
| *Ministral-3-3B-Instruct* | 0.646 | 0.546–0.734 | 0.292 | 0.844 |
| *Ministral-3-14B-Instruct* | 0.698 | 0.600–0.781 | 0.401 | 0.896 |
| *Ministral-3-14B-Reasoning* | 0.719 | 0.622–0.799 | 0.457 | 0.990 |
| *Llama-3.1-8B-Instruct* | 0.604 | 0.504–0.696 | 0.245 | 0.969 |

A similar trend was observed on the held-out test set. GPT-OSS-20B again achieved the highest raw accuracy, reaching 0.833 using Docling and 0.885 using PyMuPDF4LLM. Gemma-4-31B-it maintained consistent performance across both conversion methods, with a raw accuracy of 0.813 using both tools, while Gemma-4-26B-A4B-it achieved raw accuracies of 0.792 using Docling and 0.823 using Pymupdf4llm. Qwen3-14B achieved a raw accuracy of 0.792 under both conversion methods. Overall, the relative ranking of models remained largely consistent between the training and test sets.

Relaxed accuracy was consistently higher than raw accuracy for most evaluated models. On the training set, the highest relaxed accuracy was achieved by Ministral-3-14B-Reasoning under PyMuPDF4LLM (0.985). Gemma-4-26B-A4B-it (0.980 Docling), Ministral-3-14B-Reasoning (0.975 Docling) and Gemma-4-26B-A4B-it (0.975 PyMuPDF4LLM) followed closely. Similar findings were observed on the test set, where Mistral-3-14B-Reasoning achieved the highest relaxed accuracy using both tools, followed by Gemma-4-26B-A4B-it (0.979).

Only small differences were observed between the two PDF conversion tools. For example, GPT-OSS-20B achieved a higher raw accuracy using PyMuPDF4LLM than Docling on the test set (0.885 vs. 0.833), however, Gemma-4-31B-it achieved identical raw accuracy (0.813) using both conversion methods. Similar minor variations were observed for the remaining models, with no conversion tool consistently outperforming the other across all evaluated models.

The 95% Wilson confidence intervals of raw accuracy showed that the closely grouped top models were not clearly separated from one another. On the Docling training set, GPT-OSS-20B (0.899, 95% CI 0.848–0.933), Gemma-4-31B-it (0.888, 0.837–0.925), Gemma-4-26B-A4B-it (0.873, 0.819–0.913), and Qwen3-14B (0.863, 0.808–0.904) had broadly overlapping intervals, while all clearly exceeded the Gemma-3 models (≈0.56, upper bounds < 0.64). Cohen’s kappa mirrored this ordering under both converters: on the Docling training set κ ranged from 0.08 (Gemma-3-27B-it) to 0.80 (GPT-OSS-20B), with GPT-OSS-20B and the Gemma-4 models again in the substantial range (κ ≈ 0.75–0.80) and Qwen3-14B close behind (κ = 0.73). Notably, the Gemma-3 models combined non-trivial raw accuracy (∼0.55–0.62) with near-zero κ (< 0.10 on Docling), indicating their agreement with human judgments was scarcely better than chance. Agreement was comparable under PyMuPDF4LLM (training κ from 0.12 to 0.76), consistent with the small overall effect of the conversion tool on screening performance.

Runtime varied substantially across models but showed relatively small differences between the two conversion methods. GPT-OSS-20B completed full-text screening in 58.13 minutes using Docling and 61.47 minutes using PyMuPDF4LLM. Gemma-4-26B-A4B-it required 22.71 and 27.42 minutes, respectively, while Gemma-4-31B-it required 43.87 and 48.83 minutes. In contrast, Ministral-3-14B-Reasoning required the longest processing time, taking 743.15 minutes with Docling and 669.71 minutes with PyMuPDF4LLM.

## Discussion

Our study demonstrated that local LLMs can effectively support article screening when integrated into a carefully designed screening workflow. We developed an end-to-end workflow that included sample dataset construction, prompt engineering, model-specific output extraction strategies, and performance validation for both abstract and full-text screening. When paired with structured input data, iteratively refined prompts, and carefully designed output processing strategies, several models achieved strong screening performance. Notably, several local LLMs achieved performance comparable to human reviewers and commercial LLMs, while offering additional advantages including local deployment, greater control over data security, flexible customization, and reduced operating costs. Based on these findings, we provide a set of practical recommendations (Table 3) to guide researchers in developing reliable and reproducible local LLM-assisted article screening workflow.

**Table 3.** Practical recommendations for using local LLMs in literature screening.

| Steps | Recommendation |
| --- | --- |
| **Input Data Preparation** | Clean and standardize screening text data before running model inference. |
| **Sample Dataset Testing** | Validate the screening workflow on a small, representative dataset pre-classified by human reviewers (ground truth) to rigorously evaluate model performance metrics before full-scale deployment. |
| **Model Selection** | Balance classification performance against operational constraints, including hardware requirements, runtime, context length, and workflow integration. |
| **Prompt Engineering** | Treat prompt design as a dynamic, iterative process. Continuously refine prompts rather than relying on a fixed, single-turn instruction. |
| **Hyperparameters and Output Extraction Strategies** | Carefully inspect model outputs and optimize both generation hyperparameters (e.g., `do_sample`, `max_token`) and output extraction strategies prior to large-scale screening. |
| **Human Oversight** | Deploy local LLMs strictly as reviewer-support tools to augment human efficiency, rather than fully autonomous screeners. |

The decision to involve LLMs in screening depends on a careful trade-off between the scale of the target literature and the initial resource investment required to build an automated workflow. In this study, the choice was driven by the complexity of our review topic. In recent years, there has been rapid growth in the application of ML methods for treatment effect estimation in medical research [33–35], creating a need for a comprehensive review to identify and evaluate the most appropriate ML methods. The literature search generated 52,498 records after deduplication. Given this scale, human screening would require substantial time and effort, which motivated us to explore the use of LLMs to support the screening. Nevertheless, although local LLMs have the potential to accelerate screening tasks, developing and validating an automated workflow requires considerable effort and time. For reviews involving a small number of records, manual screening may be more efficient. For larger reviews, however, the initial investment in a sample dataset would be worthwhile to assess a workflow of using LLM to improve screening performance and efficiency. Researchers should therefore consider the size of the review, available human and computational resources, and expected screening burden when deciding whether to use an LLM-assisted workflow.

Previous studies have reported human abstract screening accuracies ranging from 79% to 94% [35]. Similar performance has also been reported for commercial LLMs such as GPT-4 [36–40]. Evidence regarding local LLMs, however, remains limited. A recent study [21] using smaller open-source models, including Mistral 7B, Vicuna-13B, and Llama 3.2, reported a best abstract screening accuracy of 66% and concluded that fully automated scoping reviews remained suboptimal. Given the rapidly expanding landscape of open-source LLMs, model selection emerged as an important factor influencing screening performance. In our study, we evaluated models from different families and generations (e.g., GPT-OSS, Llama, Gemma3, Gemma4), parameter sizes (Gemma-3-12B-it, Gemma-3-27B-it; Qwen3-4B, Qwen3-14B), and model types (instruction-tuned and thinking/reasoning-oriented) to better understand how these characteristics affect article screening. We observed that screening performance varied considerably across models. While GPT-OSS-20B and Gemma-4-31B-it achieved the strongest overall performance (raw accuracy above 87%, relaxed accuracy above 96%), smaller models generally provided faster inference and required fewer computational resources. Reasoning models typically had longer inference times due to token expended on chain of thought reasoning before a final answer, which is also reflected in our study with Ministral-3-14B-Reasoning-2512 had the longest inference time (10.09 hours for abstract screening on the training set). Although direct comparisons should be interpreted cautiously because of differences in review topics and study design, our findings suggest that local LLMs can support abstract screening at a level approaching human performance. They also indicate that high screening performance is not limited to proprietary models but can also be achieved using new generations of local LLMs, which offer additional advantages including enhanced data security, more flexible workflow customization, and cost savings.

Full-text screening improved raw accuracy while reducing false positives comparing to abstract screening, suggesting that the inclusion of full-text content provides additional information for eligibility assessment, despite the increased computational overhead. The relative ranking of models remained largely unchanged, with GPT-OSS-20B, Gemma-4-31B-it, Gemma-4-26B-A4B-it, and Qwen3-14B consistently performing strongest. The choice of PDF-to-text conversion tool had no significant impact on screening performance. Additionally, our experiments revealed that while increasing the character budget initially improved screening accuracy, performance plateaued once the text length exceeded approximately 40,000 characters (around 6,500 words). This indicates that simply expanding the context window does not guarantee better context processing, and striking the right balance between computational efficiency and content preservation remains a challenge. Consequently, future work should explore more granular full-text processing techniques, such as section-specific extraction or semantic chunking, to maximize informational value while minimizing the context window. Runtime varied substantially across models, again highlighting the importance of trade-off between computational efficiency and screening performance. Overall, our findings suggest that selecting a model for literature screening should involve balancing screening performance with practical considerations such as hardware requirements, inference speed, and ease of integration into the workflow.

Using a sample dataset and implementing a rigorous train-test split were essential to enable robust internal validation. We found it valuable to first develop and evaluate the workflow using a representative sample dataset. Sample testing allows researchers to assess workflow feasibility, identify potential issues, optimize prompts and output extraction strategies, and estimate computational requirements before processing large numbers of records. Importantly, the sample dataset should adequately represent all screening categories used in the study (e.g., Relevant, Not Relevant, and Uncertain). Within each category, articles should represent a variety of scenarios to ensure that model performance is assessed across diverse screening situations. Furthermore, to support workflow development and evaluation, screening records should be maintained in a structured and standardized format. For abstract screening, useful fields include article titles, abstracts, human classifications (ground truth), and the rationale underlying human screening decisions. Recording screening rationales can facilitate error analysis, help explain disagreements between human reviewers and LLMs, and support prompt refinement. Missing information, formatting errors, and document conversion issues will affect model predictions and performance assessment. Therefore, screening records should be carefully cleaned and standardized before model inference and evaluation.

Beyond model selection and data preparation, prompt design and output processing played important roles in ensuring reliable and reproducible LLM-assisted screening workflows. In our study, prompt development was an iterative process, and screening performance improved as the prompt was progressively refined based on misclassified records and the corresponding model reasoning in the training data. Effort was invested in defining concepts that could be interpreted inconsistently by LLMs, such as “AI methods” and “treatment effects”. To obtain reliable screening decisions, prompts should clearly specify the screening task, eligibility criteria, classification rules, and desired output format, while potentially ambiguous concepts should be explicitly defined. We also observed no improvement from few-shot prompting compared with zero-shot prompting, consistent with previous findings [30]. Output processing requires careful consideration as well. We observed large variation in response formats across models, with some placing classifications at the beginning of the response, others in the middle or at the end. Models may also generate reasoning that truncates the final classification. To address these issues, several extraction strategies were designed, including extracting classifications from different sections of model responses and increasing the maximum token limit when necessary. We additionally used greedy decoding (`do_sample=False)` and “`return_full_text=False`” to improve reproducibility and simplify output processing. Together, these findings highlight the importance of validating model outputs and optimizing both generation settings and extraction strategies before large-scale deployment.

Human oversight remains essential throughout the screening workflow. Several inherent limitations of LLMs were observed in our study, highlighting the need for human verification. Models relied on contextual assumptions rather than explicit evidence. For example, some models inferred the presence of medical datasets from general discussions of causal inference or healthcare applications even when such information was not clearly specified. This usually resulted in false “Relevant” classifications. Another common source of error was distinguishing predictive modeling from treatment effect estimation. LLMs frequently misclassified studies focused on risk prediction or prognostic modeling as studies that estimated treatment effect, despite explicit instructions in the prompt. We also observed inconsistencies between model reasoning and final classifications, with some models correctly identifying unmet eligibility criteria but ultimately assigning an “Uncertain” or “Relevant” classification rather than “Not Relevant.” As a result, disagreements between model and human classifications persisted despite overall strong performance. These findings highlight the continued importance of human oversight, particularly for borderline cases. Taken together, these findings suggest that local LLMs currently function most effectively as reviewer-support tools that can accelerate screening workflows rather than fully autonomous reviewers.

Several directions for future research may further advance the use of local LLMs in evidence synthesis. First, model performance was evaluated using a sample dataset from a single review topic. Applying the workflow to complete review datasets from diverse research domains would provide a more realistic assessment of its practical value and efficiency. More research is needed to evaluate the generalizability of the screening workflow and to build empirical rules for the recommended sample dataset sizes. Second, future studies may extend the application of local LLMs beyond literature screening to other stages of the review process, such as literature searching, data extraction, and meta-analysis. Our study focused only on the screening stage; integrating local LLMs across multiple review tasks may further reduce the workload associated with evidence synthesis. Furthermore, future work should also focus on improving screening precision. In our sample screening, a substantial proportion of irrelevant studies (i.e., false positives) were incorrectly classified as “Relevant”, resulting in considerable manual effort during later review stages. Developing a second-stage screening prompt or multi-stage screening strategy may help reduce false positives and the manual workload required to exclude them in the later stages. Lastly, future work could explore LLM fine-tuning on human-annotated datasets or evaluating higher-parameter local models, provided sufficient computational resources are available.

## Acknowledgements

We gratefully acknowledge Dr. Alisa Berger for her expert support in designing and conducting the systematic literature search.

## Contributions

LL and HC built and annotated the dataset as human reviewers, designed the screening strategy, analyzed the data, and wrote the original draft. LL, HC, HGY, and MAP conceptualized the study. WS validated the results and supported data analysis. WS and NS provided the computational resources. MAP and NS supervised the project. All authors reviewed and approved the final manuscript.

## Trial Registration

Clinical trial number: not applicable.

## Data Availability

All the data analysis in this paper was conducted using Python version 3.14. The code used for the analyses conducted in this study is available at Github (https://github.com/hongrchen/LLM4SLR).

## Ethics declarations

Ethics approval and consent to participate

Not applicable.

Consent for publication

Not applicable.

## Figures and Tables

## Supplementary Materials

### S1. Literature Search

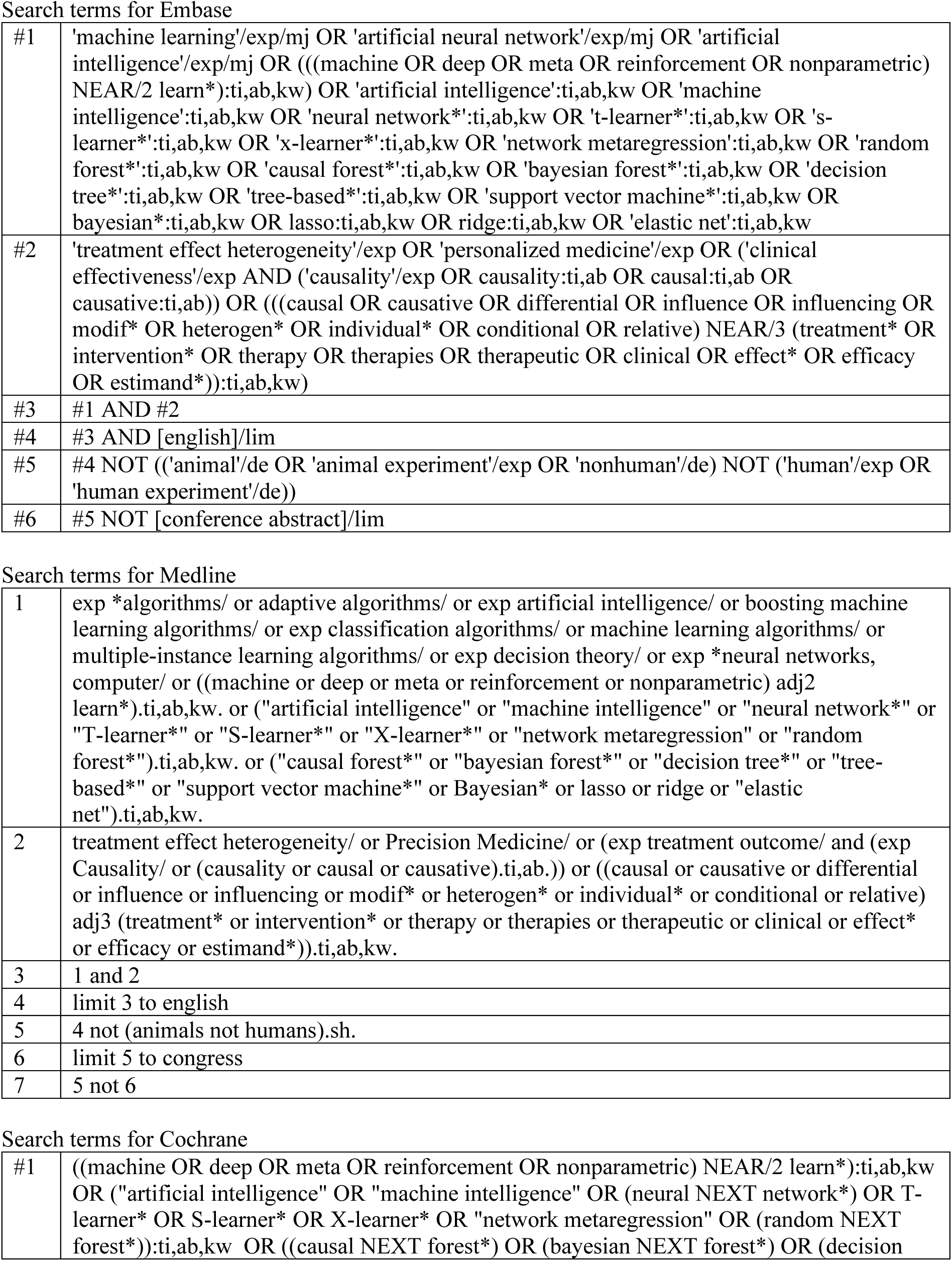
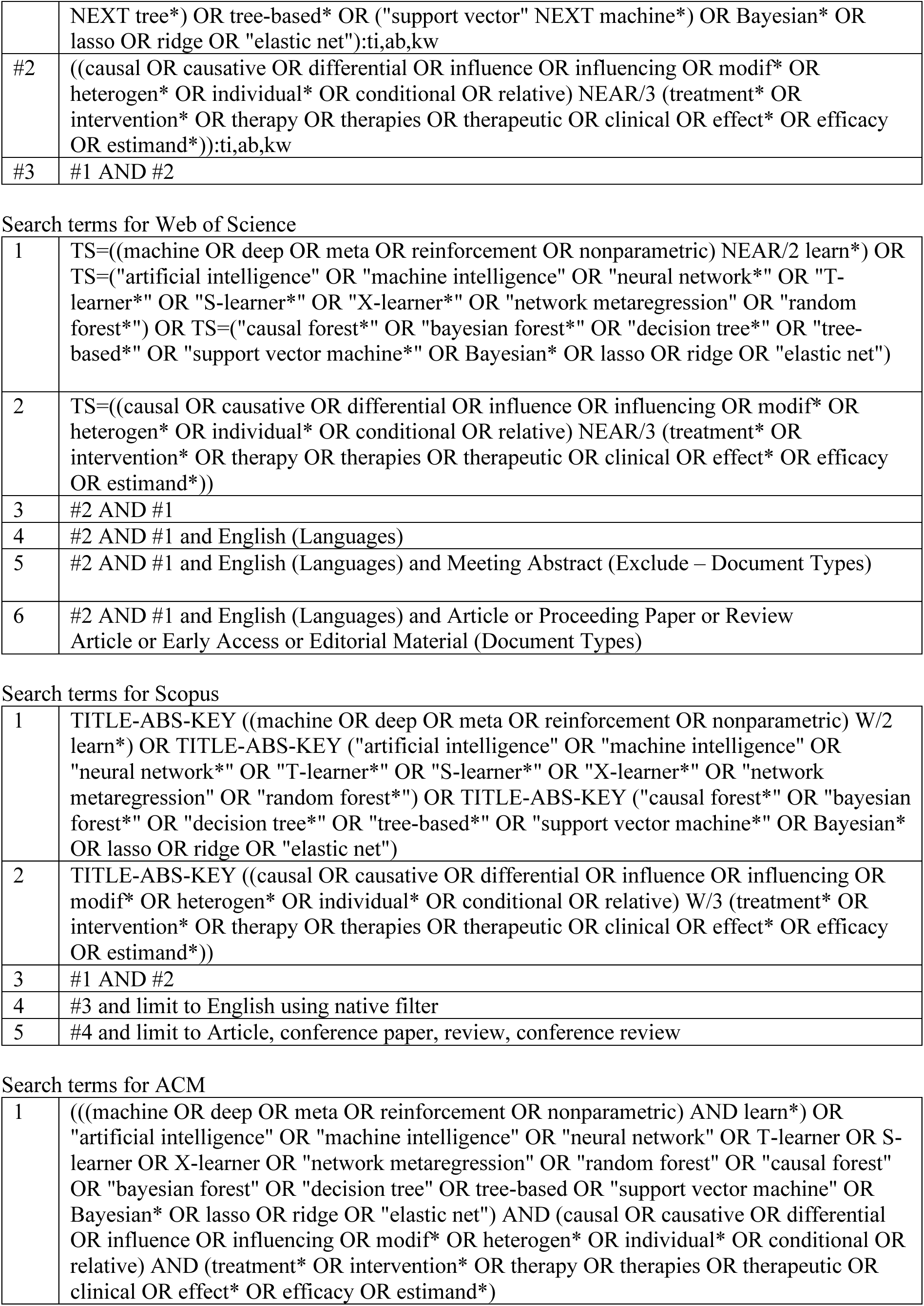

Search terms for Embase
|  |  |
| --- | --- |
| #1 | 'machine learning'/exp/mj OR 'artificial neural network'/exp/mj OR 'artificial intelligence'/exp/mj OR (((machine OR deep OR meta OR reinforcement OR nonparametric) NEAR/2 learn*):ti,ab,kw) OR 'artificial intelligence':ti,ab,kw OR 'machine intelligence':ti,ab,kw OR 'neural network*':ti,ab,kw OR 't-learner*':ti,ab,kw OR 's-learner*':ti,ab,kw OR 'x-learner*':ti,ab,kw OR 'network metaregression':ti,ab,kw OR 'random forest*':ti,ab,kw OR 'causal forest*':ti,ab,kw OR 'bayesian forest*':ti,ab,kw OR 'decision tree*':ti,ab,kw OR 'tree-based*':ti,ab,kw OR 'support vector machine*':ti,ab,kw OR 'bayesian*':ti,ab,kw OR lasso:ti,ab,kw OR ridge:ti,ab,kw OR 'elastic net':ti,ab,kw |
| #2 | 'treatment effect heterogeneity'/exp OR 'personalized medicine'/exp OR ('clinical effectiveness'/exp AND ('causality'/exp OR causality:ti,ab OR causal:ti,ab OR causative:ti,ab)) OR (((causal OR causative OR differential OR influence OR influencing OR modif* OR heterogen* OR individual* OR conditional OR relative) NEAR/3 (treatment* OR intervention* OR therapy OR therapies OR therapeutic OR clinical OR effect* OR efficacy OR estimand*)):ti,ab,kw) |
| #3 | #1 AND #2 |
| #4 | #3 AND [english]/lim |
| #5 | #4 NOT (('animal'/de OR 'animal experiment'/exp OR 'nonhuman'/de) NOT ('human'/exp OR 'human experiment'/de)) |
| #6 | #5 NOT [conference abstract]/lim |
| Search terms for Medline |  |
| 1 | exp *algorithms/ or adaptive algorithms/ or exp artificial intelligence/ or boosting machine learning algorithms/ or exp classification algorithms/ or machine learning algorithms/ or multiple-instance learning algorithms/ or exp decision theory/ or exp *neural networks, computer/ or ((machine or deep or meta or reinforcement or nonparametric) adj2 learn*).ti,ab,kw. or ("artificial intelligence" or "machine intelligence" or "neural network*" or "T-learner*" or "S-learner*" or "X-learner*" or "network metaregression" or "random forest*").ti,ab,kw. or ("causal forest*" or "bayesian forest*" or "decision tree*" or "tree-based*" or "support vector machine*" or Bayesian* or lasso or ridge or "elastic net").ti,ab,kw. |
| 2 | treatment effect heterogeneity/ or Precision Medicine/ or (exp treatment outcome/ and (exp Causality/ or (causality or causal or causative).ti,ab.)) or ((causal or causative or differential or influence or influencing or modif* or heterogen* or individual* or conditional or relative) adj3 (treatment* or intervention* or therapy or therapies or therapeutic or clinical or effect* or efficacy or estimand*)).ti,ab,kw. |
| 3 | 1 and 2 |
| 4 | limit 3 to english |
| 5 | 4 not (animals not humans).sh. |
| 6 | limit 5 to congress |
| 7 | 5 not 6 |
| Search terms for Cochrane |  |
| #1 | ((machine OR deep OR meta OR reinforcement OR nonparametric) NEAR/2 learn*):ti,ab,kw OR ("artificial intelligence" OR "machine intelligence" OR (neural NEXT network*) OR T-learner* OR S-learner* OR X-learner* OR "network metaregression" OR (random NEXT forest*)):ti,ab,kw OR ((causal NEXT forest*) OR (bayesian NEXT forest*) OR (decision |
|  | NEXT tree*) OR tree-based* OR ("support vector" NEXT machine*) OR Bayesian* OR lasso OR ridge OR "elastic net"):ti,ab,kw |
| #2 | ((causal OR causative OR differential OR influence OR influencing OR modif* OR heterogen* OR individual* OR conditional OR relative) NEAR/3 (treatment* OR intervention* OR therapy OR therapies OR therapeutic OR clinical OR effect* OR efficacy OR estimand*)):ti,ab,kw |
| #3 | #1 AND #2 |
| Search terms for Web of Science |  |
| 1 | TS=((machine OR deep OR meta OR reinforcement OR nonparametric) NEAR/2 learn*) OR TS=("artificial intelligence" OR "machine intelligence" OR "neural network*" OR "T-learner*" OR "S-learner*" OR "X-learner*" OR "network metaregression" OR "random forest*") OR TS=("causal forest*" OR "bayesian forest*" OR "decision tree*" OR "tree-based*" OR "support vector machine*" OR Bayesian* OR lasso OR ridge OR "elastic net") |
| 2 | TS=((causal OR causative OR differential OR influence OR influencing OR modif* OR heterogen* OR individual* OR conditional OR relative) NEAR/3 (treatment* OR intervention* OR therapy OR therapies OR therapeutic OR clinical OR effect* OR efficacy OR estimand*)) |
| 3 | #2 AND #1 |
| 4 | #2 AND #1 and English (Languages) |
| 5 | #2 AND #1 and English (Languages) and Meeting Abstract (Exclude – Document Types) |
| 6 | #2 AND #1 and English (Languages) and Article or Proceeding Paper or Review Article or Early Access or Editorial Material (Document Types) |
| Search terms for Scopus |  |
| 1 | TITLE-ABS-KEY ((machine OR deep OR meta OR reinforcement OR nonparametric) W/2 learn*) OR TITLE-ABS-KEY ("artificial intelligence" OR "machine intelligence" OR "neural network*" OR "T-learner*" OR "S-learner*" OR "X-learner*" OR "network metaregression" OR "random forest*") OR TITLE-ABS-KEY ("causal forest*" OR "bayesian forest*" OR "decision tree*" OR "tree-based*" OR "support vector machine*" OR Bayesian* OR lasso OR ridge OR "elastic net") |
| 2 | TITLE-ABS-KEY ((causal OR causative OR differential OR influence OR influencing OR modif* OR heterogen* OR individual* OR conditional OR relative) W/3 (treatment* OR intervention* OR therapy OR therapies OR therapeutic OR clinical OR effect* OR efficacy OR estimand*)) |
| 3 | #1 AND #2 |
| 4 | #3 and limit to English using native filter |
| 5 | #4 and limit to Article, conference paper, review, conference review |
| Search terms for ACM |  |
| 1 | ((((machine OR deep OR meta OR reinforcement OR nonparametric) AND learn*) OR "artificial intelligence" OR "machine intelligence" OR "neural network" OR T-learner OR S-learner OR X-learner OR "network metaregression" OR "random forest" OR "causal forest" OR "bayesian forest" OR "decision tree" OR tree-based OR "support vector machine" OR Bayesian* OR lasso OR ridge OR "elastic net") AND (causal OR causative OR differential OR influence OR influencing OR modif* OR heterogen* OR individual* OR conditional OR relative) AND (treatment* OR intervention* OR therapy OR therapies OR therapeutic OR clinical OR effect* OR efficacy OR estimand*)) |

### S2. Summary of Model Characteristics

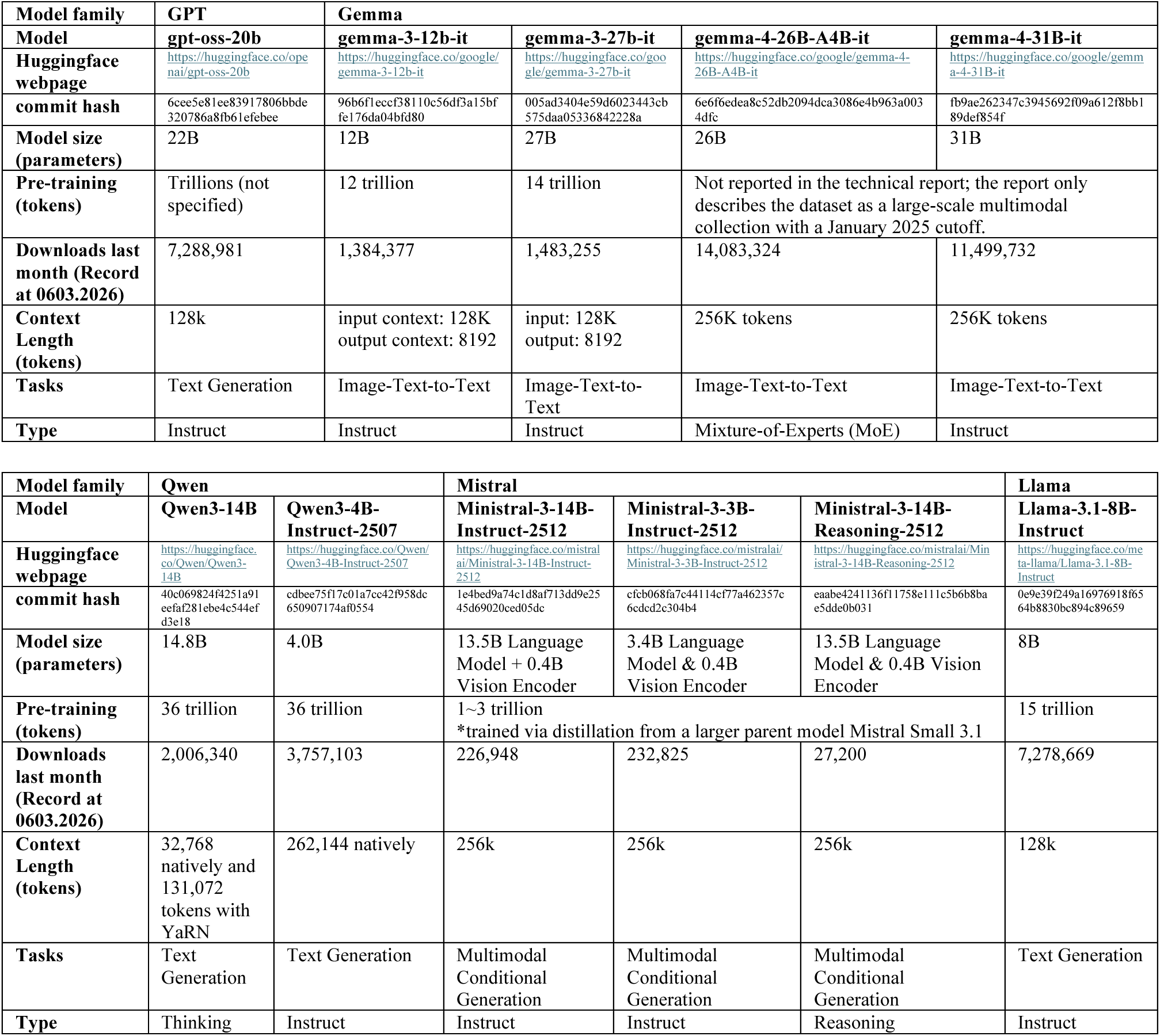

### S3. Prompt for Title and Abstract Screening

The screening prompt was developed through iterative refinement using the training data.

“““You are reviewing texts to determine its relevance based on predefined criteria. Carefully read the provided texts and evaluate whether it meets each of the criteria below. Base your judgment only on the information provided in the texts. If information is missing or unclear, do not assume—use the “Uncertain” category when appropriate.

We define the following three criteria:

1. Use of AI Methods The study must use an algorithmic learning approach rather than a fixed statistical formula. A method counts as AI if it meets at least one of the following: it automatically models non-linear relationships and interactions (e.g., Trees, Neural Nets, Bayesian models); or it uses algorithmic optimization like Boosting, Bagging, or Cross-Validated tuning; or it uses regularization (like Lasso) to handle high-dimensional variable sets. Standard “Fixed-Effect” models (OLS, Logistic Regression, 2-Stage Least Squares) and standard Hypothesis Tests (T-tests, ANOVA) are not AI.
2. Estimation of Treatment Effects The study must attempt to answer a “What if?” question (Counterfactual effect such as ATE, CATE, HTE, ITE). It must measure the impact of an intervention on an outcome using AI methods. Exclude purely prognostic/predictive models. For example, a model that predicts “Risk of Heart Attack” is not a treatment effect study unless it specifically measures how a specific medication changes that risk.
3. Use of Real-World Medical Data The study must use real-world medical data (e.g., observational data or clinical trial data related to medical interventions or health outcomes), and the AI methods used for estimating treatment effects must be applied to this data.

Classification Rules:

Classify as “Relevant” if all three criteria are clearly satisfied.

Classify as “Not Relevant” if any one of the three criteria is clearly not satisfied.

Classify as “Uncertain” if at least one criterion is unclear, and the remaining criteria are satisfied. For example, classify as Uncertain if the methodology or data source is not mentioned, or if the medical nature of the data is ambiguous. However, if the context clearly implies a non-medical domain or a purely statistical (non-AI) approach, classify as Not Relevant.

Output Format:

Your response must be exactly one of the following: “Relevant”, “Not Relevant”, “Uncertain”.

Now evaluate the following article:

Title: {title}

Abstract: {abstract}”””

## References

1. Snyder H. Literature review as a research methodology: An overview and guidelines. Journal of Business Research. 2019;104:333–9. 10.1016/j.jbusres.2019.07.039.

2. Sucharew H, Macaluso M. Methods for Research Evidence Synthesis: The Scoping Review Approach. Journal of Hospital Medicine. 2019;14:416–8. 10.12788/jhm.3248.

3. Arksey H, O’Malley L. Scoping studies: towards a methodological framework. International Journal of Social Research Methodology. 2005;8:19–32. 10.1080/1364557032000119616.

4. Chen H, Jiang Z, Liu X, Xue CC, Yew SME, Sheng B, et al. Can large language models fully automate or partially assist paper selection in systematic reviews? British Journal of Ophthalmology. 2025;109:962–6. 10.1136/bjo-2024-326254.

5. Ge L, Agrawal R, Singer M, Kannapiran P, De Castro Molina JA, Teow KL, et al. Leveraging artificial intelligence to enhance systematic reviews in health research: advanced tools and challenges. Syst Rev. 2024;13:269. 10.1186/s13643-024-02682-2.

6. Blaizot A, Veettil SK, Saidoung P, Moreno-Garcia CF, Wiratunga N, Aceves-Martins M, et al. Using artificial intelligence methods for systematic review in health sciences: A systematic review. Research Synthesis Methods. 2022;13:353–62. 10.1002/jrsm.1553.

7. Khraisha Q, Put S, Kappenberg J, Warraitch A, Hadfield K. Can large language models replace humans in systematic reviews? Evaluating GPT-4’s efficacy in screening and extracting data from peer-reviewed and grey literature in multiple languages. Research Synthesis Methods. 2024;15:616–26. 10.1002/jrsm.1715.

8. Borah R, Brown AW, Capers PL, Kaiser KA. Analysis of the time and workers needed to conduct systematic reviews of medical interventions using data from the PROSPERO registry. BMJ Open. 2017;7:e012545. 10.1136/bmjopen-2016-012545.

9. Guo E, Gupta M, Deng J, Park Y-J, Paget M, Naugler C. Automated Paper Screening for Clinical Reviews Using Large Language Models: Data Analysis Study. Journal of Medical Internet Research. 2024;26:e48996. 10.2196/48996.

10. Gates A, Gates M, DaRosa D, Elliott SA, Pillay J, Rahman S, et al. Decoding semi-automated title-abstract screening: findings from a convenience sample of reviews. Syst Rev. 2020;9:272. 10.1186/s13643-020-01528-x.

11. Polanin JR, Pigott TD, Espelage DL, Grotpeter JK. Best practice guidelines for abstract screening large-evidence systematic reviews and meta-analyses. Res Synth Methods. 2019;10:330–42. 10.1002/jrsm.1354.

12. Thode L, Iftikhar U, Mendez D. Exploring the use of LLMs for the selection phase in systematic literature studies. Information and Software Technology. 2025;184:107757. 10.1016/j.infsof.2025.107757.

13. Ofori-Boateng R, Aceves-Martins M, Wiratunga N, Moreno-Garcia CF. Towards the automation of systematic reviews using natural language processing, machine learning, and deep learning: a comprehensive review. Artif Intell Rev. 2024;57:200. 10.1007/s10462-024-10844-w.

14. Sundaram G, Berleant D. Automating Systematic Literature Reviews with Natural Language Processing and Text Mining: a Systematic Literature Review. 2023. 10.48550/arXiv.2211.15397.

15. van de Schoot R, de Bruin J, Schram R, Zahedi P, de Boer J, Weijdema F, et al. An open source machine learning framework for efficient and transparent systematic reviews. Nat Mach Intell. 2021;3:125–33. 10.1038/s42256-020-00287-7.

16. Ouzzani M, Hammady H, Fedorowicz Z, Elmagarmid A. Rayyan—a web and mobile app for systematic reviews. Syst Rev. 2016;5:210. 10.1186/s13643-016-0384-4.

17. Li M, Sun J, Tan X. Evaluating the effectiveness of large language models in abstract screening: a comparative analysis. Syst Rev. 2024;13:219. 10.1186/s13643-024-02609-x.

18. Syriani E, David I, Kumar G. Assessing the Ability of ChatGPT to Screen Articles for Systematic Reviews. Journal of Computer Languages. 2024;80:101287. 10.1016/j.cola.2024.101287.

19. Tran V-T, Gartlehner G, Yaacoub S, Boutron I, Schwingshackl L, Stadelmaier J, et al. Sensitivity, specificity and avoidable workload of using a large language models for title and abstract screening in systematic reviews and meta-analyses. 2023;:2023.12.15.23300018. 10.1101/2023.12.15.23300018.

20. Singhal K, Azizi S, Tu T, Mahdavi SS, Wei J, Chung HW, et al. Large language models encode clinical knowledge. Nature. 2023;620:172–80. 10.1038/s41586-023-06291-2.

21. Bayani A, Epoh Ewane LP, Oliveira dos Anjos DS, Mac-Seing M, Nikiema JN. Leveraging open-source large language models (LLMs) in scoping reviews: a case study on disability and AI applications. International Journal of Medical Informatics. 2025;204:106048. 10.1016/j.ijmedinf.2025.106048.

22. Zhang G, Jin Q, Zhou Y, Wang S, Idnay B, Luo Y, et al. Closing the gap between open source and commercial large language models for medical evidence summarization. npj Digit Med. 2024;7:239. 10.1038/s41746-024-01239-w.

23. Savage CH, Kanhere A, Parekh V, Langlotz CP, Joshi A, Huang H, et al. Open-Source Large Language Models in Radiology: A Review and Tutorial for Practical Research and Clinical Deployment. Radiology. 2025;314:e241073. 10.1148/radiol.241073.

24. Builtjes L, Bosma J, Prokop M, van Ginneken B, Hering A. Leveraging open-source large language models for clinical information extraction in resource-constrained settings. Jamia Open. 2025;8:ooaf109. 10.1093/jamiaopen/ooaf109.

25. Kochetkov D. Who Owns the Knowledge? Copyright, GenAI, and the Future of Academic Publishing. 2026. 10.48550/arXiv.2511.21755.

26. Balloccu S, Schmidtová P, Lango M, Dušek O. Leak, Cheat, Repeat: Data Contamination and Evaluation Malpractices in Closed-Source LLMs. 2024. 10.48550/arXiv.2402.03927.

27. Manchanda J, Boettcher L, Westphalen M, Jasser J. The Open Source Advantage in Large Language Models (LLMs). 2025. 10.48550/arXiv.2412.12004.

28. Kukreja S, Kumar T, Purohit A, Dasgupta A, Guha D. A Literature Survey on Open Source Large Language Models. In: Proceedings of the 2024 7th International Conference on Computers in Management and Business. New York, NY, USA: Association for Computing Machinery; 2024. p. 133–43. 10.1145/3647782.3647803.

29. Galli C, Gavrilova AV, Calciolari E. Large Language Models in Systematic Review Screening: Opportunities, Challenges, and Methodological Considerations. Information. 2025;16:378. 10.3390/info16050378.

30. Syriani E, David I, Kumar G. Screening articles for systematic reviews with ChatGPT. Journal of Computer Languages. 2024;80:101287. 10.1016/j.cola.2024.101287.

31. Livathinos N, Auer C, Lysak M, Nassar A, Dolfi M, Vagenas P, et al. Docling: An Efficient Open-Source Toolkit for AI-driven Document Conversion. 2025. 10.48550/arXiv.2501.17887.

32. pymupdf/pymupdf4llm. https://github.com/pymupdf/pymupdf4llm. 2026.

33. Sanchez P, Voisey JP, Xia T, Watson HI, O’Neil AQ, Tsaftaris SA. Causal machine learning for healthcare and precision medicine. R Soc Open Sci. 2022;9:220638. 10.1098/rsos.220638.

34. Emmenegger C, Spohn M-L, Elmer T, Bühlmann P. Treatment Effect Estimation with Observational Network Data using Machine Learning. Journal of Causal Inference. 2025;13:20230082. 10.1515/jci-2023-0082.

35. Moccia C, Moirano G, Popovic M, Pizzi C, Fariselli P, Richiardi L, et al. Machine learning in causal inference for epidemiology. Eur J Epidemiol. 2024;39:1097–108. 10.1007/s10654-024-01173-x.

36. Ali F, Tan AS-C, Wang SJ-W. Can machine learning help accelerate article screening for systematic reviews? Yes, when article separability in embedding space is high. Research Synthesis Methods. 2025;16:194–210. 10.1017/rsm.2024.16.

37. Sanghera R, Thirunavukarasu AJ, El Khoury M, O’Logbon J, Chen Y, Watt A, et al. High-performance automated abstract screening with large language model ensembles. J Am Med Inform Assoc. 2025;32:893–904. 10.1093/jamia/ocaf050.

38. Ramchandani R, Guo E, Rakab E, Rathod J, Strain J, Klement W, et al. Validation of automated paper screening for esophagectomy systematic review using large language models. PeerJ Comput Sci. 2025;11:e2822. 10.7717/peerj-cs.2822.

39. Zuo C, Yang X, Errickson J, Li J, Hong Y, Wang R. AI-assisted evidence screening method for systematic reviews in environmental research: integrating ChatGPT with domain knowledge. Environ Evid. 2025;14:5. 10.1186/s13750-025-00358-5.

40. Insuk S, Boonpattharatthiti K, Booncharoen C, Chaipitak P, Rashid M, Veettil SK, et al. How Well Do ChatGPT and Claude Perform in Study Selection for Systematic Review in Obstetrics. J Med Syst. 2025;49:110. 10.1007/s10916-025-02246-4.

